# Cardiomyocyte Globotriaosylceramide Accumulation in Adult Male Patients with Fabry Disease and IVS4 + 919G>A GLA Mutation is Progressive with Age and Correlates with Left Ventricular Hypertrophy and Reduced Left Ventricular Ejection Fraction

**DOI:** 10.1101/2023.12.09.23298489

**Authors:** Fu-Pang Chang, Ting-Rong Hsu, Sheng-Che Hung, Shih-Hsien Sung, Wen-Chung Yu, Dau-Ming Niu, Behzad Najafian

**Affiliations:** Institute of Clinical Medicine, National Yang Ming Chiao Tung University, Taipei, Taiwan; Department of Pathology and Laboratory Medicine, Taipei Veterans General Hospital, Taipei, Taiwan; Department of Pediatrics, Taipei Veterans General Hospital, Taipei, Taiwan; Division of Neuroradiology, Department of Radiology, University of North Carolina Chapel Hill, North Carolina, USA; Biomedical Research Imaging Center, University of North Carolina Chapel Hill, North Carolina, USA; Department of Internal Medicine, College of Medicine, National Yang Ming Chiao Tung University, Taipei, Taiwan; Division of Cardiology, Department of Medicine, Taipei Veterans General Hospital, Taipei, Taiwan; Department of Laboratory Medicine & Pathology, University of Washington, Seattle, USA

**Keywords:** Fabry, cardiomyopathy, globotriaosylceramide, stereology

## Abstract

**Background:** While cardiovascular complications are the most common cause of mortality in Fabry disease, the relationship between globotriaosylceramide (GL-3) accumulation, the hallmark of Fabry cardiomyopathy, and cardiac hypertrophy has not been fully elucidated.

**Methods:** We developed unbiased stereology protocols to quantify the ultrastrcture of Fabry cardiomyopathy. Endomyocardial biopsies from 10 adult male enzyme replacement therapy naïve Fabry patients with IVS4 + 919G>A GLA mutation were studied. The findings were correlated with cardiac MRI and clinical data.

**Results:** Ultrastructural parameters showed significant relationships with key imaging and clinical and functional variables. Average cardiomyocyte volume and GL-3 volume per cardiomyocyte were progressively increased with age. Eighty-four percent of left ventricular mass index (LVMI) variability was explained by cardiomyocyte nuclear volume, age and plasma globotriaosylsphingosine with cardiomyocyte nuclear volume being the only independent predictor of LVMI. Septal thickness was directly and left ventricular ejection fraction (LVEF) was inversely correlated with cardiomyocyte GL-3 accumulation. Sixty-five percent of left ventricular ejection fraction (LVEF) variability was explained by cardiomyocyte GL3 volume, serum α-galactosidase-A activity and age with cardiomyocyte GL3 volume being the only independent predictor of LVEF. Residual α-galactosidase-A activity was directly correlated with myocardial microvasculature density.

**Conclusions:** The unbiased stereological methods introduced in this study unraveled novel relationships between cardiomyocyte structure and important imaging and clinical parameters. These novel tools can help better understand Fabry cardiomyopathy pathophysiology.

**Clinical Perspective:** *What is new?:*

- We developed a novel unbiased quantitative approach to study cardiac biopsies by electron microscopy.
- Ultrastructural parameters showed significant relationships with key clinical, imaging, cardiac functional variables in cardiac biopsies from patients with Fabry disease.

*What are the clinical implications?:*

- These quantitative approaches are powerful tools to better understand pathophysiology of Fabry disease and potentially in other cardiomyopathies.

## Introduction

Fabry disease (MIM 301500), an X-linked lysosomal storage disorder, is caused by mutations in GLA gene leading to deficiency of the enzyme α-galactosidase-A (α-Gal-A) and an inability to catabolize neutral glycosphingolipids, particularly globotriaosylceramide (GL3)(1). The progressive accumulation of GL3 in many organs results in a wide range of clinical manifestations.

There are two major phenotypes of Fabry disease: “classic” and “later-onset” variants(2). In classic Fabry males, no or very low α-Gal-A activity is associated with extensive accumulation of GL3 in various cell types including endothelial cells, and manifests with characteristic skin lesions (angiokeratoma), ocular lesions (corneal and lenticular opacities), neuropathic pains in the extremities (acroparesthesia), and gastrointestinal symptoms during childhood or adolescence; whereas, in later-onset Fabry disease, presence of residual α-Gal-A activity is associated with no or low GL3 accumulation in endothelial cells, lack of early manifestations in classic phenotype, and delayed incidence of serious complications by later ages(2). Female patients have more variable presentations due to random X-inactivation(3). While cardiovascular complications are the number one cause of mortality in Fabry disease, cardiac biopsy studies are scarce and no validated systematic methodology to study cardiac biopsies from patients with Fabry disease have been proposed(4). Subsequently, relationships between cardiac pathology and function have not been elucidated. We have developed unbiased stereology methods to quantify GL3 content of glomerular cells at electron microscopy details(5–7). These studies have been instrumental in unraveling relationships between age and GL3 accumulation, injury and loss of podocytes, which play pivotal role in glomerular structure and function(5). Podocytes similar to cardiomyocytes are post-mitotic cells and show resistance to enzyme replacement therapy (ERT)(8, 9). Importantly, our unbiased quantitative approaches allowed us to document partial clearance of podocytes from GL3 accumulation following ERT or chaperone therapy(6, 10, 11).

In this work, we adapted similar methodology to study the structure of cardiomyocytes in biopsies from treatment-naïve male Fabry patients with later-onset/cardiac variant due to IVS4+919G>A GLA mutation which is mainly associated with cardiac manifestations including hypertrophic cardiomyopathy, and arrhythmias later in life(12, 13). This study, for the first time, demonstrates relationships between cardiomyocyte GL3 accumulation, left ventricular hypertrophy and cardiac function.

## Methods

### Subjects

The study protocol has been approved by the Institutional Review Board of Taipei Veterans General Hospital, Taipei, Taiwan (IRB no. 2015-11-008AC). Informed consent was waived due to the retrospective nature of the study. Ten treatment-naïve male Fabry patients carrying the later-onset IVS4 + 919G>A mutation from Taipei Veterans General Hospital in Taiwan were enrolled in this study. The patients’ demographic data, laboratory results and imaging findings were extracted from medical records. GLA mutation, plasma or leukocyte α-Gal-A enzyme activity, plasma lyso-GL3 and plasma GL3 levels were analyzed as described in the previous publication(14, 15). α-Gal-A activity was measured in 8/10 patients in plasma samples and in 2/10 in leukocytes. Leukocyte and plasma α-Gal-A activity levels are strongly correlated(16). In order to make leukocyte and plasma α-Gal-A activity values measured at different times comparable, we expressed the enzyme activity as a percentage of the laboratory’s mean of normal population(5).

### Cardiac Imaging Studies

All patients underwent gadolinium-enhanced cardiac magnetic resonance imaging using a 3.0-T scanner (Discovery MR750, GE Healthcare, Milwaukee, Wisconsin). Left ventricular mass index (LVMI) was calculated as left ventricular mass normalized to height^2.7^, and left ventricular hypertrophy (LVH) was defined by an LVMI of >48 g/m^2.7^ in men(17, 18). Interventricular septal thickness was also measured. Myocardial late gadolinium enhancement (LGE) was evaluated at 10 min after intravenous administration of 0.1 mmol/ kg gadobutrol on a breath-hold electrocardiogram (ECG)-gated 2-dimensional myocardial delayed enhancement sequence (slice thickness, 8mm; both short-axis and 4-chamber views; field of view, 340 × 316 mm; matrix, 256 × 192) and the number of affected LV segments were recorded (13, 19).

### Endomyocardial Biopsies

Cardiac biopsies from treatment-naïve patients with Fabry disease were performed as part of the Taiwan treatment guidelines for confirmation of Fabry cardiomyopathy when applying for ERT funding from the National Health Insurance. Two biopsies from non-Fabry patients with arrhythmia were included as controls. No diagnostic changes can be identified by routine histopathological examinations in these two controls. Catheterization was performed through right internal jugular vein and under digital X-ray guidance. Endomyocardial bioptome was inserted into the right ventricle, and specimens were obtained from the interventricular septum.

### Electron Microscopy Studies

Tissues were fixed in 2.5% glutaraldehyde in phosphate buffer, post-fixed with 1% osmium tetroxide in Sorenson’s phosphate buffer, followed by dehydration through a graded series of ethanol washes, and embedded in Spurr’s Epon. Semithin (1 µm) sections were cut from the block and stained with toluidine blue for microscopic review. Thin sections were mounted on Formvar-coated slot copper grids and examined using a JEOL 1230 transmission EM. Overlapping digital images (∼2,000X magnification) were taken with an AMT XR80 camera. Montages of the entire section profiles were constructed using Adobe Photoshop software (Adobe Photoshop CS5 Extended, version 12.0.4 x64) for stereological quantitative studies.

### Stereological Studies

All visible CM nuclei were identified and numbered in the entire section profile montages. Average volume of CM nuclei [V(CMN)] was estimated using the point-sampled intercept (PSI) method with slight modification to minimize the volume-weighted property of this method, as detailed elsewhere for similar quantitative studies for podocytes(6). This method provides an unbiased shape-independent estimation of the volume of complex-shaped particles. A visual description of the protocol is provided in **Figure 1**. Briefly, a point grid was superimposed on each nucleus and a random point was selected using a random number generator. Assuming that CM nuclei are positioned in random directions, a horizontal and a vertical line were passed though the sampling point as random direction lines. If all nuclei are convex, an unbiased estimate of their average volume 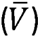 will be 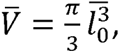 where 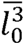 is the average of the 3^rd^ power of the lengths of intercept of the random line and nuclear membrane passing through the sampling point. If the nuclear shape includes concavities, the random direction line may create more of the average nuclear volume will be 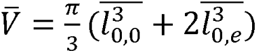, where *l*_0,0_ is the distance between the immediate intercepts of the random line with the nuclear membrane on both sides of the sampling point. 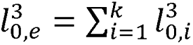 or sum of 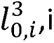 over all the extra intercepts not containing the sample point, while 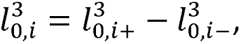 where 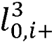 and 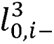 are the longest and the shortest distances between the sampling point and intercepts of the random direction line and nuclear membrane.

**Figure 1.**
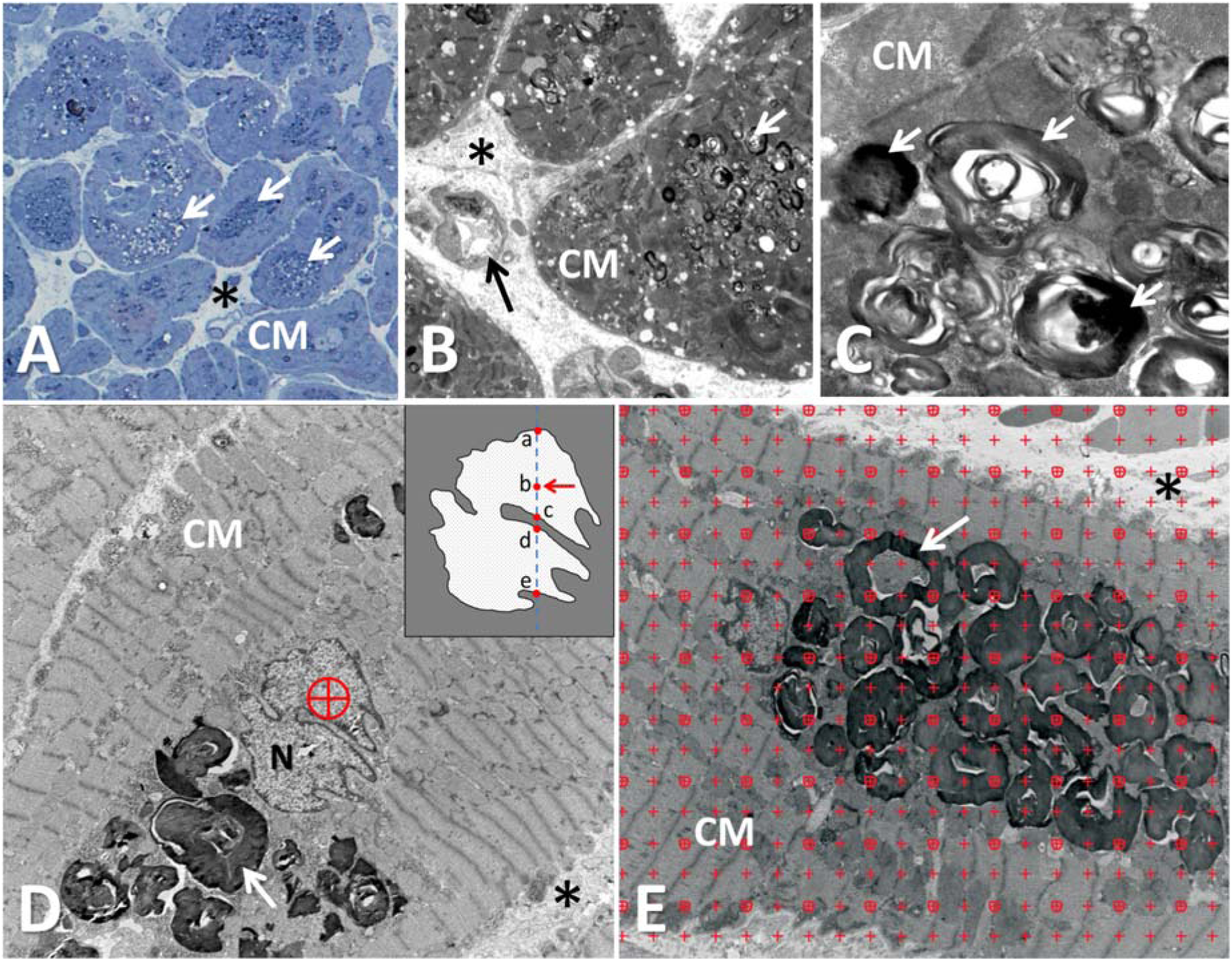
Endomyocardial biopsy from a patient with Fabry disease. **(A)** A semi-thin (1 um) section stained with toluidine blue shows accumulation of cardiomyocytes with numerous dark round inclusions. **(B)** A low magnification transmission electron microscopy image of the biopsy. The black arrow shows an interstitial capillary; the asterisk shows interstitium. **(C)** a higher magnification view shows a cardiomyocyte (CM) with many globotriaosylceramide (GL3) inclusions (white arrows). **(D)** A random point (red circled cross) was selected on the nucleus (N) of a cardiac myocyte. A schematic view of the nucleus is shown in the inset. The intersections of a vertical line (dashed line) passing through the random point (point b shown by the red arrow) mark the points on the nuclear membrane for the following measurement needed for the point-sampled intercept method (see text) : *l_0,0_*=*ac; l_0,i+_*=*be*; and *l_0,i_*_-_=*bd*. **(E)** a point grid superimposed on cardiomyocytes (CM) for estimation of Vv(GL3/CM) (see text); white arrow shows GL3 inclusions; the asterisk shows interstitium.

In order to identify the minimum number of nuclear profiles needed to be measured to obtain a robust and stable estimate of the average V(CMN), incremental sampling with five nuclei per step was performed (**Supplemental Figure 1**). Thus, stabilization of V(CMN) values was observed after inclusion of sixty nuclei. Therefore, sixty nuclei were used as the minimum requirement for a representative modified PSI measurement. If the initial section profile in one given case contained less than sixty nuclei, an additional deeper section profile separated by 10 μm was prepared to obtain more nuclei. Section profiles of these 10 cases contained a variable number of CMN (71 [60-167], median [range]).

Volume fraction of CMN per CM [Vv(CMN/CM)], volume fraction of GL3 inclusions per CM [Vv(Inc/CM)], volume fraction of blood vessels in cardiac parenchyma [Vv(Ves/Heart)], and volume fraction of interstitial compartment in cardiac parenchyma [Vv(Int/Heart)] were estimated by point counting method (**Figure 1E**). Then, average CM volume [V(CM)] was calculated as: *V(CM)=V(CMN)/Vv(CMN/CM)*. The total volume of GL3 inclusions per CM [V(Inc/CM)] which represents the volume of GL3 inclusions per CM independent of cellular enlargement or shrinkage was calculated as: *V(Inc/CM)=V(CM).Vv(Inc/CM)*.

By electron microscopy, both GL3 inclusions and lipofuscin granules are electron dense (i.e. dark) and usually accumulate around the nuclei. While GL3 inclusions are typically multilamellar and different from lipofuscin granules which are solid black with partially grey areas in the middle, it is difficult to tell these structures apart when they are mixed together (**Suppl. Figure 2**). Therefore, when point counting method was performed on section profiles from Fabry patients, the points that either fell on typical GL3 inclusions (multilamellar myelin figures and zebra bodies) or solid dark granules (indistinguishable from lipofuscin) were included in the estimation of Vv(Inc/CM). The volume density of lipofuscin granules per CM and total volume of lipofuscin granules per CM in two non-Fabry control biopsies were provided for reference.

### Statistical analysis

Statistica 13 (TIBCO Software Inc) was used for statistical analysis. Graphpad prism and Microsoft Excel were used to generate the graphs. Variables were expressed as mean ± SD or median [range] depending on their type (parametric or non-parametric) or distribution. Due to limited number of subjects, association between the variables were examined by Spearman rank order correlation. General linear model was used to adjust for confounding variables. Multiple regression analysis was used to explore the contribution of multiple predictor variables on a dependent variable. P<0.05 was considered statistically significant.

## Results

The clinical characteristics of the Fabry patients are summarized in **Table 1**. All patients were adults, age 62[44-68] (median [range]). Low residual α-Gal-A enzyme activity was present in all patients and ranged from 7.79 to 25.21% (median=12.19%) of the laboratory’s mean of normal population. Plasma lyso-GL3 levels were elevated in 9/10 and borderline in one subject (5.20[2.56-10.4] nM; laboratory normal<2.6). Plasma GL3 concentration was 5.14[1.70-9.69] µg/mL (laboratory normal < 5.7). Half (5/10) of the patients had increased left ventricular mass index (LVMI), 54.1[32.4-139.6] g/m^2.7^ (normal for men<48(17)). However, left ventricular ejection fraction (LVEF) was normal in all patients, 73[57-79]% (normal≥57%)(20). Late gadolinium enhancement (LGE) in MRI was present in 9/10 patients. LVMI and septal thickness were strongly correlated (r=0.93, p=0.0001).

**Table 1.** Clinical and imaging characteristics of 10 treatment-naïve male Fabry patients with later-onset IVS4 + 919G>A mutation.

| | Age | Gender | α-Gal-A | Lyso-GL3 | GL3 | eGFR | LVMI | Septal thickness (mm) | LVEF (%) | LGE |
| --- | --- | --- | --- | --- | --- | --- | --- | --- | --- | --- |
| 1 | 40s | M | 15.52 | 3.50 | 5.20 | 72.70 | 68.14 | 25.9 | 71 | 3 |
| 2 | 40s | M | 13.20 | 2.56 | 7.20 | 64.92 | 32.44 | 12.7 | 79 | 7 |
| 3 | 50s | M | 12.37 | 3.21 | 5.08 | 93.64 | 41.22 | 13.0 | 79 | 0 |
| 4 | 60s | M | 25.21 | 6.51 | 9.69 | 89.19 | 46.18 | 14.2 | 72 | 3 |
| 5 | 60s | M | 12.28 | 5.60 | 2.60 | 74.43 | 43.39 | 19.8 | 78 | 1 |
| 6 | 60s | M | 10.17 | 4.80 | 3.50 | 77.78 | 47.68 | 17.0 | 62 | 1 |
| 7 | 60s | M | 11.05 | 10.40 | 6.39 | 98.09 | 139.58 | 30.0 | 75 | 7 |
| 8 | 60s | M | 12.10 | 7.47 | 5.44 | 79.96 | 98.85 | 30.7 | 57 | 12 |
| 9 | 60s | M | 11.82 | 7.13 | 2.66 | 41.24 | 106.37 | 27.0 | 67 | 5 |
| 10 | 60s | M | 7.79 | 4.30 | 1.70 | 38.83 | 60.48 | 22.2 | 73 | 2 |
| Ctrl 1 | 40s | M | N/A | N/A | N/A | 104.56 | 38.50 | 10.0 | 65 | 0 |
| Ctrl 2 | 60s | M | N/A | N/A | N/A | 69.89 | 39.20 | 10.0 | 60 | 0 |

**Abbreviations:** α-Gal-A = α-galactosidase-A activity expressed as a percentage of the laboratory's mean of normal population; Lyso-GL3 = plasma globotriaosylsphingosine level (reference ranges: <2.6 nM); GL-3 = plasma globotriaosylceramide level (reference ranges: <5.7 ug/ml); eGFR = estimated glomerular filtration rate (normal value: >60 ml/min/1.73 m<sup>2</sup>); LVMI = Left ventricular mass index (normal value < 48 g/m<sup>2.7</sup> in men); LVEF = left ventricular ejection fraction; LGE = late gadolinium enhancement expressed as the number of affected LV segments ; M = male; N/A = not available.

### IVS4+919G>A GLA mutation is associated with characteristic features of Fabry cardiomyopathy

The electron microscopic studies showed variable amounts of GL-3 inclusions accumulated in the cytoplasm of cardiomyocytes in all cardiac biopsies from Fabry patients. The GL3 inclusions were characterized by round to ovoid electron-dense multi-lamellar structures (**Figure 1**). The distribution of GL3 inclusions in cardiomyocytes was typically clustered around the nuclei. In more severely affected cases, the massive intracytoplasmic GL3 inclusions displaced the cardiac sarcomere. Rare GL3 inclusions were present in the interstitial tissue, but no inclusions were found in capillary endothelial cells, consistent with the later-onset Fabry phenotype.

### Cardiomyocyte hypertrophy and GL3 accumulation progressively increase with age in Fabry disease

Structural parameters measured in the cardiac biopsies are shown in **Table 2**. While due to the limited number of control cases no meaningful statistical comparison could be done between Fabry and control endomyocradial biopsies, the average V(CM) in Fabry biopsies (49,917±30,419 µm^3^) was ∼1.9 fold greater than control V(CM) values (24,803 and 28,685 µm^3^), consistent with cardiomyocyte hypertrophy. Likewise, average V(CMN) in Fabry biopsies (266±145 µm^3^) was ∼1.9 fold greater than control V(CMN) values (138 and 142 µm^3^), consistent with nuclear enlargement, a feature that is often attributed to cellular injury, activation or hypertrophy. The nucleocytoplasmic ratio (proportion between nuclear volume and cytoplasmic volume) of cardiomyocytes in Fabry biopsies (0.006±0.003) remained quite close to that in controls (0.005 in both). While lipofuscin granules, which can be present in normal cardiomyocytes, do not have a multilamellated appearance which is typical of GL3 inclusions. Since on random profiles it was difficult to readily distinguish between these two types of material in Fabry biopsies, they were combined in quantitative studies and considered “inclusions”. Thus, cardiomyocyte GL3/inclusion volume fraction was ∼33 fold greater in Fabry (0.06 ± 0.03) than in control biopsies (0.007 and 0.005). However, cardiomyocyte GL3 volume was 3,193 ± 2378 µm^3^ in Fabry biopsies. In other words, only ∼6% [1.7 - 10.2%] of the cardiomyocyte volume was composed of GL-3 inclusions and the observed cellular enlargement was mainly due to hypertrophy of cytoplasmic compartments other than GL3 inclusions. On the other hand, cardiomyocyte GL-3 volume and volume of cardiomyocyte cytoplasm which was not occupied by GL-3 inclusions [V((CM-Inc)/CM)] were strongly correlated [r=0.92, p=0.0002].

**Table 2.** Structural parameters in cardiac biopsies from Fabry patients (cases 1-10) and two controls (Ctrl 1 and 2).

|  | V(CMN) | Vv(CMN/CM) | Vv(Inc/CM) | V(CM) | V(Inc/CM) | V((CM-Inc)/CM) | Vv(Int/Heart) | Vv(Ves/Heart) | Nv(CM/Heart) |
| --- | --- | --- | --- | --- | --- | --- | --- | --- | --- |
| 1 | 133.08 | 0.0064 | 0.0704 | 20949 | 1474 | 19475 | 0.1933 | 0.0200 | 4.1370E-5 |
| 2 | 133.10 | 0.0046 | 0.0367 | 28828 | 1058 | 27770 | 0.1158 | 0.0337 | 2.9211E-5 |
| 3 | 150.71 | 0.0050 | 0.0212 | 30294 | 643 | 29650 | 0.1333 | 0.0465 | 2.7342E-5 |
| 4 | 125.93 | 0.0105 | 0.0170 | 11956 | 204 | 11752 | 0.1445 | 0.0526 | 6.8988E-5 |
| 5 | 253.35 | 0.0044 | 0.0606 | 57663 | 3496 | 54168 | 0.0796 | 0.0184 | 1.5572E-5 |
| 6 | 260.19 | 0.0041 | 0.0870 | 63087 | 5492 | 57595 | 0.1225 | 0.0127 | 1.3730E-5 |
| 7 | 530.30 | 0.0118 | 0.0499 | 45093 | 2252 | 42841 | 0.0873 | 0.0267 | 1.8854E-5 |
| 8 | 396.40 | 0.0035 | 0.0588 | 112386 | 6609 | 105778 | 0.0700 | 0.0375 | 7.8969E-6 |
| 9 | 445.51 | 0.0054 | 0.0721 | 82285 | 5935 | 76350 | 0.0624 | 0.0178 | 1.0837E-5 |
| 10 | 230.90 | 0.0050 | 0.1022 | 46628 | 4766 | 41862 | 0.1348 | 0.0191 | 1.7832E-5 |
| Ctrl 1 | 133.70 | 0.0054 | 0.0053† | 24803 | 132* | 24671 | 0.1051 | 0.0124 | 3.6051E-5 |
| Ctrl 2 | 141.59 | 0.0049 | 0.0069† | 28685 | 198* | 28487 | 0.0902 | 0.0157 | 3.2127E-5 |

**Abbreviations:** [V(CMN)] = average volume of cardiomyocyte nuclei (CMN); [Vv(CMN/CM)] = volume fraction of CMN per cardiomyocyte (CM); [Vv(Inc/CM)] = volume fraction of GL3 inclusions per CM; [V(CM)] = average CM volume; [V(Inc/CM)] = total volume of GL3 inclusions per CM; [V((CM-Inc)/CM)] = volume of cardiomyocyte cytoplasm which was not occupied by GL3 inclusions; [Vv(Int/Heart)] = volume fraction of interstitial compartment in cardiac parenchyma; [Vv(Ves/Heart)] = volume fraction of blood vessels in cardiac parenchyma; [Nv(CM/heart)] = cardiomyocyte number density
†: volume fraction of lipofuscin granules per CM in non-Fabry controls
\*: total volume of lipofuscin granules per CM in non-Fabry controls

Importantly, age was directly associated with multiple structural parameters related to cardiomyocyte size or GL3 content in patients with Fabry disease, including average cardiomyocyte volume (r=0.73, p=0.02), volume of cardiomyocyte nuclei (r=0.71, p=0.02), cardiomyocyte GL3 volume [V(Inc/CM)] (r=0.74, p=0.02) and cardiomyocyte volume which was not GL3 inclusions (r=0.69, p=0.03) (**Figure 2**). These results are indicative that cardiomyocyte hypertrophy and GL3 accumulation are both progressive with age in this cardiac/later-onset variant of Fabry disease. Overall, the relationships between age and biopsy structural variables related to Fabry cardiomyopathy were stronger than those with MRI parameters. Thus, LVMI and septal thickness showed direct trends of association with age (r=0.57, p=0.08; and r=0.60, p=0.07, respectively), and there was no relationship between age and LVEF or LGE (**Figure 2**).

**Figure 2.**
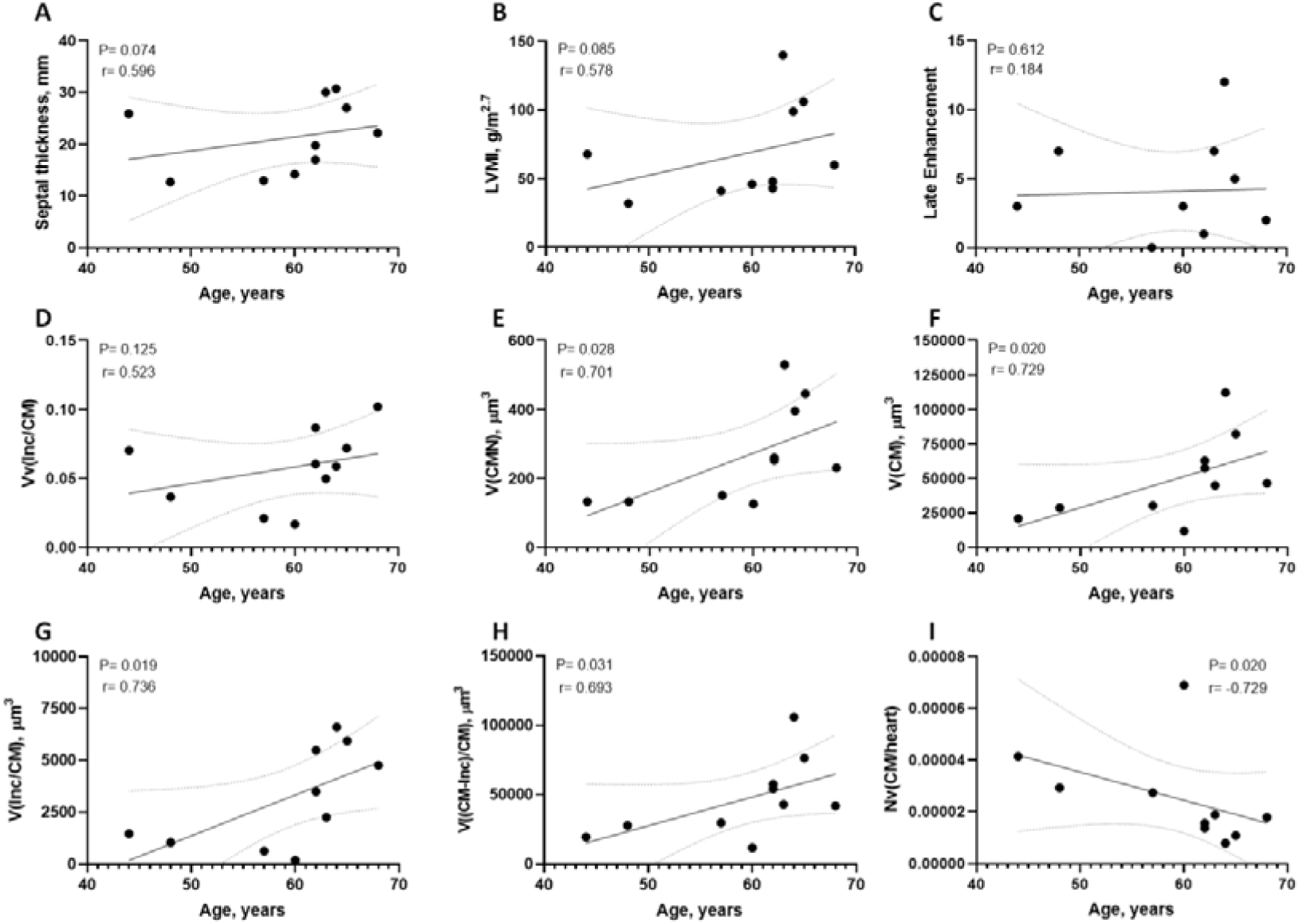
Associations between age and MRI and biopsy parameters of Fabry cardiomyopathy. Age showed **(A)** trend of direct association with septal thickness, **(B)** trend of direct association with left ventricular mass index (LVMI), **(C)** no relationship with late Gadolinium enhancement, **(D)** no relationship with cardiomyocyte GL3 volume fraction [Vv(Inc/CM)], **(E)** direct relationship with average volume of cardiomyocyte nuclei [V(CMN)], **(F)** direct relationship with average volume of cardiomyocytes [V(CM)], **(G)** direct relationship with cardiomyocyte GL3 volume [V(Inc/CM)]; **(H)** direct relationship with volume of cardiomyocyte cytoplasm which

### Correlations between MRI and biopsy indices of Fabry cardiomyopathy and left ventricular ejection fraction

Among the cardiac MRI parameters, LVMI showed direct relationship with volume of cardiomyocyte nuclei (r=0.66, p=0.04) and a trend of direct association with cardiomyocyte GL3 volume (r=0.58, p=0.08), and septal thickness was directly correlated with cardiomyocyte nuclear volume (r=0.67, p=0.03) and cardiomyocyte GL3 volume (r=0.69, p=0.03) (**Figure 3**). In order to explore factors associated with LVH, we performed forward stepwise multiple regression analysis with LVMI as the dependent variable and age, α-Gal-A activity, plasma lyso-GL3, and stereological parameters as predictors. The model explained 84% (adjusted R^2^, p=0.002) of LVMI variability. Cardiomyocyte nuclear volume, age and lyo-GL3 were the only variables remaining in the model with cardiomyocyte nuclear volume being the only statistically significant (p=0.02) independent predictor of LVMI.

**Figure 3.**
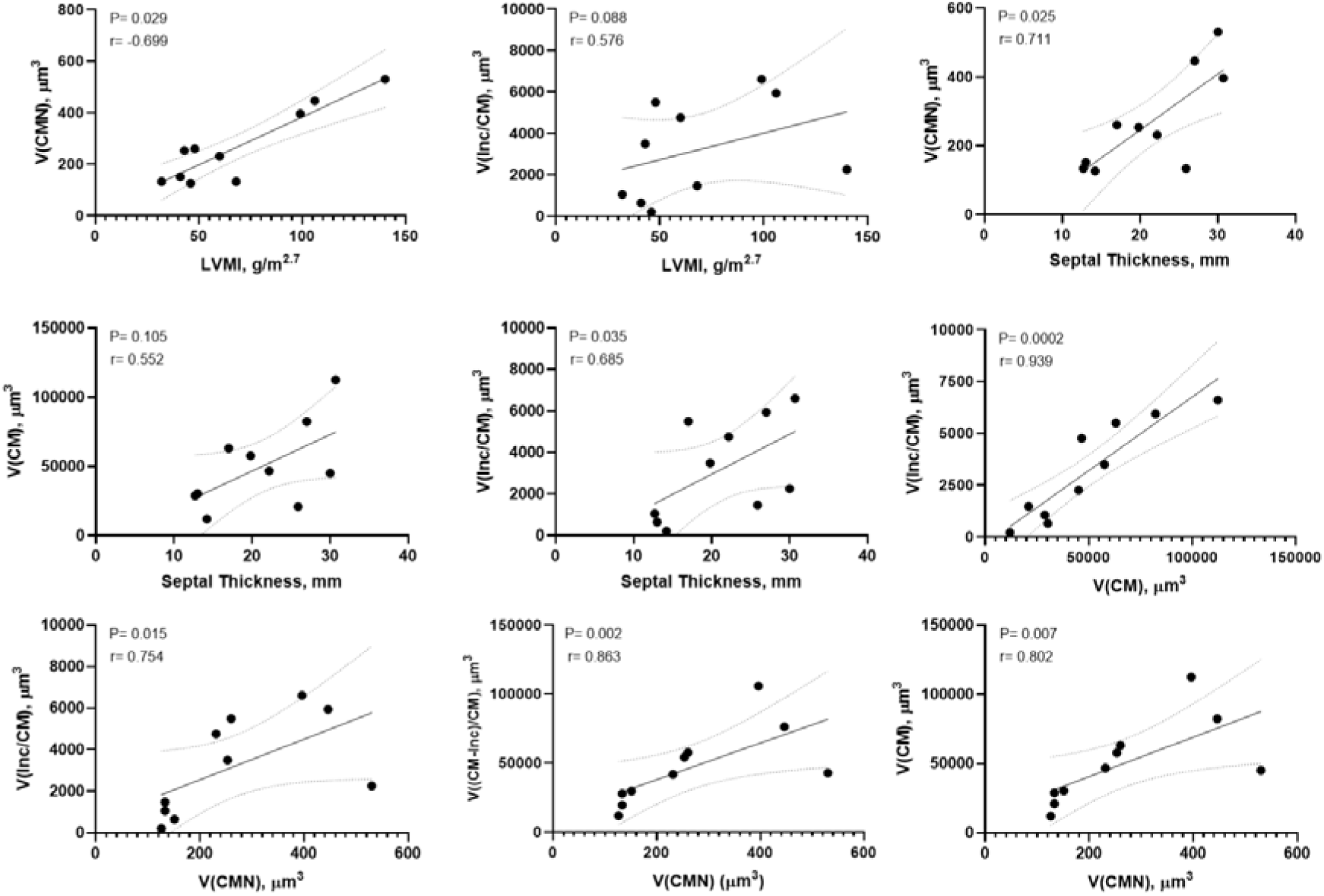
Correlations between MRI and biopsy indices of Fabry cardiomyopathy. **(A-B)** Left ventricular mass index (LVMI) showed **(A)** direct correlation with cardiomyocyte nuclear volume [V(CMN)], and **(B)** direct correlation with cardiomyocyte GL3 volume [V(Inc/CM)]. (C-E) Septal thickness was directly related to **(C)** cardiomyocyte nuclear volume [V(CMN)], **(D)** cardiomyocyte volume V(CM), **(E)** cardiomyocyte GL3 volume [V(Inc/CM)]. **(F)** cardiomyocyte volume was directly related to cardiomyocyte GL3 volume. **(G-I)** cardiomyocyte nuclear volume was directly related to **(G)** cardiomyocyte GL3 volume, cardiomyocyte volume which is not GL3 [V((CM-Inc)/CM)] and **(I)** cardiomyocyte volume.

In addition, while MRI LVMI and septal thickness showed trends of inverse associations with LVEF, the inverse association between LVEF and cardiomyocyte GL3 volume was stronger and statistically significant (r=-0.68, p=0.036) (**Figure 4**). In order to explore factors affecting LVEF, forward stepwise multiple regression analysis with LVEF as the dependent variable and age, α-Gal-A activity, plasma lyso-GL3, LVMI and stereological parameters as predictors was performed. Sixty-five percent (adjusted R^2^, p=0.03) of LVEF variability was explained by cardiomyocyte GL3 volume, α-Gal-A activity and age. However, cardiomyocyte GL3 volume was the only statistically significant independent predictor of LVEF (p=0.007). There was also an inverse relationship between cardiomyocyte GL3 volume fraction and eGFR (r=-0.66, p=0.044) (**Figure 4**). Even though the volume of cardiomyocyte which is not GL3 had a much greater contribution than GL3 inclusions in increased cardiomyocyte volume (see above), this parameter showed no correlation with LVMI, α-Gal-A activity, lyso-GL3 or LVEF and only showed a trend of association with septal thickness (r=0.59, p=0.07).

**Figure 4.**
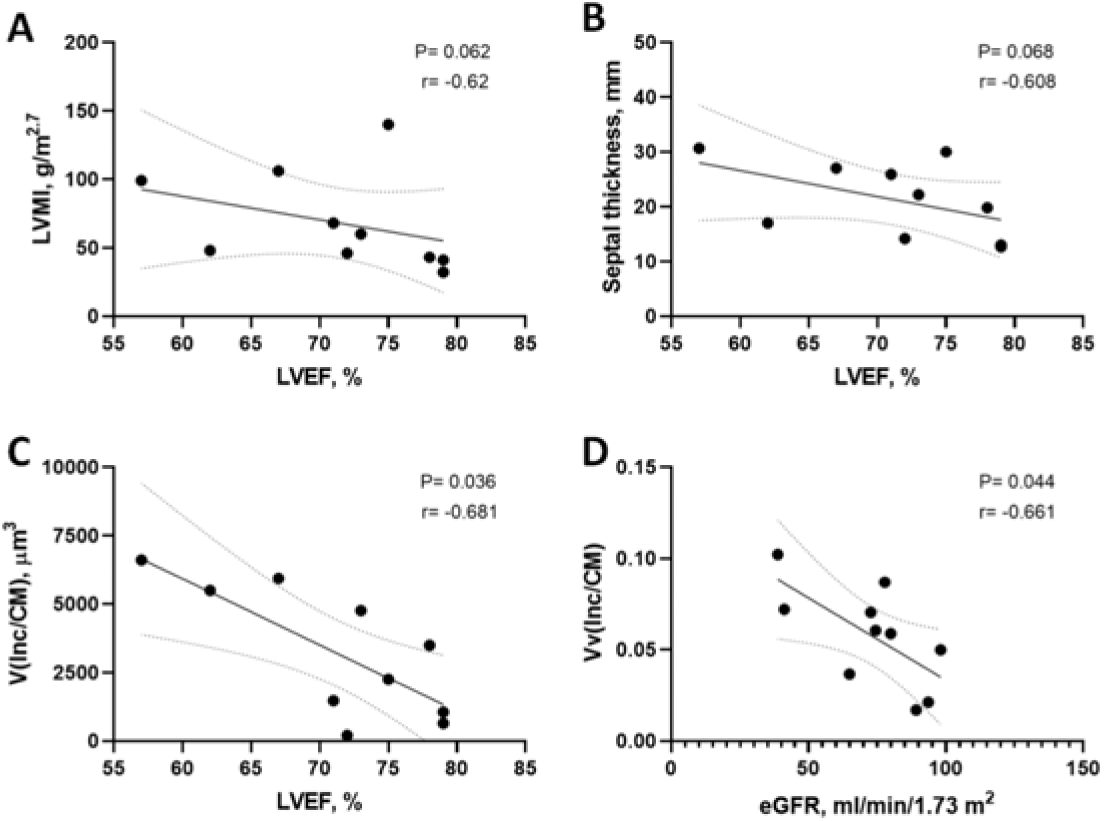
Associations of MRI and biopsy indices of Fabry cardiomyopathy with cardiac and renal function. Left ventricular ejection fraction (LVEF) showed a trend of inverse relationship with MRI **(A)** left ventricular mass index (LVMI), and **(B)** septal thickness, but statistically significant inverse relationship with **(C)** cardiomyocyte GL3 volume [V(Inc/CM)]. **(D)** There was an inverse relationship between cardiomyocyte GL3 volume fraction [Vv(Inc/CM)] and estimated glomerular filtration rate (eGFR).

### Correlations of MRI and biopsy indices of Fabry cardiomyopathy with α-Gal-A activity and plasma lyso-GL3

Residual α-Gal-A activity showed an inverse trend of association with LVMI (r=-0.60, p=0.07), but strong inverse relationship with cardiomyocyte GL3 volume fraction (r=-0.84, p=0.002), and a trend of inverse association with cardiomyocyte GL3 volume (r=-0.59, p=0.07) (**Figure 5**). In addition, residual α-Gal-A activity directly correlated with the volume fraction of microvasculature in cardiac parenchyma [Vv(Ves/Heart)] (r=0.67, p=0.03).

**Figure 5.**
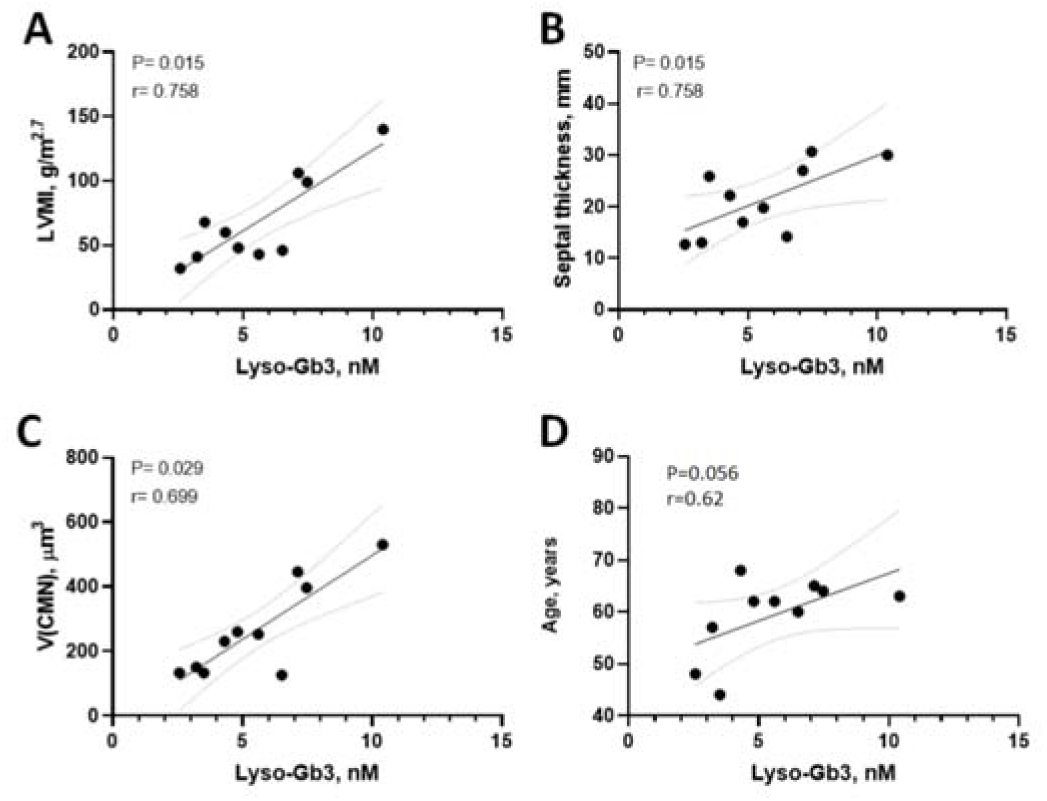
Relationships between residual α-Gal-A activity and MRI and biopsy structural parameters. α-Gal-A activity (expressed as %min. normal laboratory level) showed **(A)** trend of inverse association with LVMI, **(B)** inverse association with cardiomyocyte GL3 volume fraction [Vv(Inc/CM), **(C)** trend of inverse association with cardiomyocyte GL3 volume [V(Inc/CM)], and **(D)** direct association with cardiac microvasculature fractional volume [Vv(Ves/heart)].

Plasma lyso-GL3, a sensitive biomarker of Fabry disease(21), showed direct correlation with LVMI (r=0.76, p=0.01) and septal thickness (r=0.76, p=0.01), and among the biopsy structural parameter with cardiomyocyte nuclear volume (r=0.69, p=0.03). Plasma lyso-GL3 was observed between volume fraction of interstitium per myocardium and any of the imaging or clinical variables.also showed a trend of direct association with age (r=0.62, p=0.056) (**Figure 6**). No relationship

**Figure 6.**
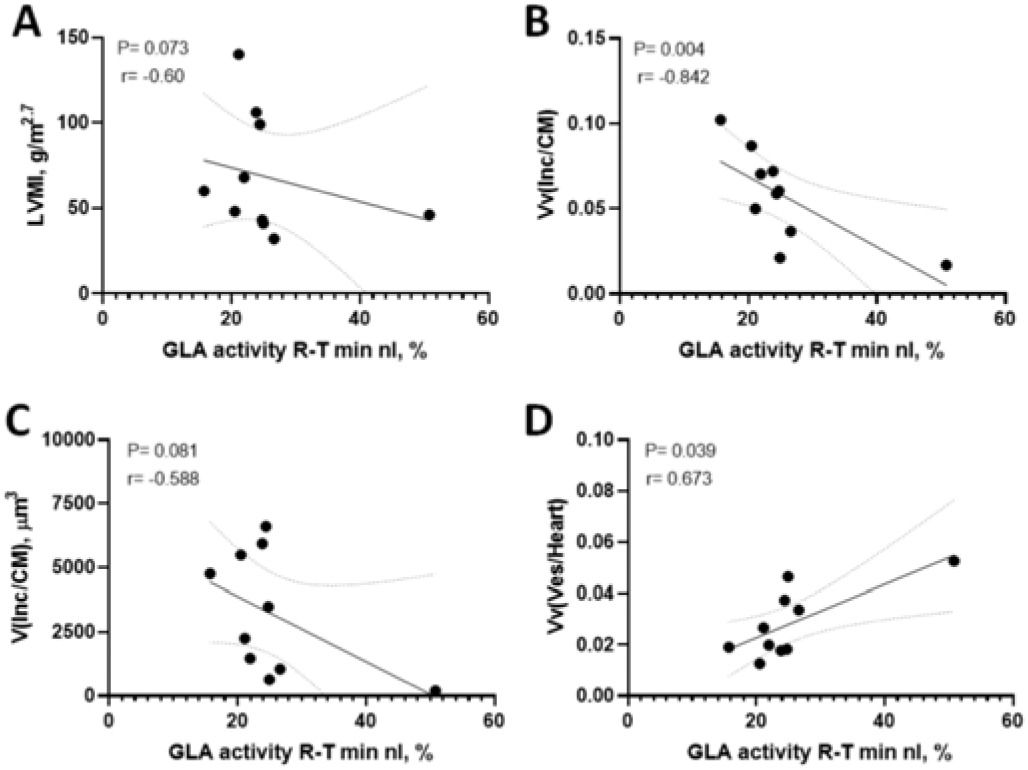
Relationships between plasma lyso-GL3, MRI, biopsy structural parameters and age. Plasma lyso-GL3 showed **(A)** direct relationship with left ventricular mass index (LVMI), **(B)** septal thickness, **(C)** cardiomyocyte nuclear volume [V(CMN)], and **(D)** age.

## Discussion

Our study offers several novel and important findings. We introduce unbiased morphometric protocols to systematically quantify ultrastructural changes of cardiomyocytes and cardiac parenchyma, including parameters specifically related to Fabry cardiomyopathy. The correlations observed between these structural parameters and clinically important variables, such as age, lyso-GL3, LVMI, septal thickness and LVEF suggest that these structural parameters can be used to better understand the pathophysiology of Fabry cardiomyopathy. In addition, accurate quantification of intracellular GL3 can inform about efficacy of Fabry specific treatments either in clinical trials or in patient’s clinical decision making.

Although cardiac involvement of Fabry disease begins early in life, the cardiac manifestations typically develop in the third to fourth decade in patients with classic disease, and present in the fourth to seventh decade in patients with late-onset phenotype (13, 22, 23). The cardiac abnormalities include myocardial hypertrophy, arrhythmias, and valvular dysfunction, while the majority of patients will present LVH with disease progression.

Our study showed that cardiomyocyte volume and cardiomyocyte GL3 volume increased with age in later-onset Fabry patients. These findings are consistent with Frustaci et al. who reported that cardiomyocyte diameter and GL3 load measured by vacuolar areas in cardiomyocyte on paraffin sections correlated with age and hypertrophy severity on cardiac imaging in classic Fabry patients(24). Cardiomyocytes are post-mitotic cells with limited regeneration capacity, therefore, the relationships between age and intracellular GL3 and cardiomyocyte volume is likely reflecting lifelong GL3 accumulation. This is similar to what had been shown for podocytes in the kidney, which are also post-mitotic(5). Thus, podocyte GL3 volume fraction linearly increases with age in young male patients with classic Fabry disease(7) and plateaus at about age 30 years, while podocyte GL3 volume (the average total GL3 volume per podocyte) keeps increasing afterward(5) and is associated with podocyte loss(5). Intracellular GL3 accumulation in cardiomyocytes, similar to podocytes(5), is associated with cellular hypertrophy. However, in contrast to podocytes where GL3 encompasses ∼30-50% of the cellular volume in classic Fabry patient, we found that only ∼6%[1.7-10.2%] of cardiomyocytes was occupied by GL3 inclusions, which is in the same scale reported by Elleder et al. in an autopsy of a patient with Fabry disease(25). This is perhaps not surprising since cardiomyocytes are highly specialized cells with non-stop vital mechanical duties which cannot afford a large fraction of their cytoplasm being occupied by inclusions. On the other hand, while both cardiomyocyte GL3 volume and cardiomyocyte volume which is not GL3 increased with age, it was only cardiomyocyte GL3 volume that was inversely correlated with LVEF and directly correlated with septal thickness, underscoring the significance of GL3 accumulation in pathogenesis of Fabry cardiomyopathy through pathways not necessarily linked to cellular hypertrophy. It is also noteworthy that all patients included in this study had LVEF values within the normal range. Thus, cardiomyocyte GL3 volume may be a more sensitive biomarker of cardiac dysfunction than LVEF itself or LVMI or septal thickness which were not associated with LVEF decline. This is reminiscent of the relationship we observed between podocytes GL3 volume fraction and urine protein creatinine ratio in young patients with Fabry disease with normal kidney function(7).

Measuring cardiomyocyte volume using unbiased stereology was critical in quantifying intracellular GL3 accumulation and unraveling the relationships between cardiomyocyte structure and other variables. The methodology introduced in this study are applicable not only to Fabry disease, but also potentially to other forms of cardiomyopathy or storage diseases affecting myocardium. In contrast to cardiomyocyte GL3 volume, cardiomyocyte GL3 volume fraction did not increase with age, likely due to cellular hypertrophy proportional to increased GL3 volume as the disease progressed. The inability to discern a change in a compartment due to proportional change in the reference space, also known as “reference trap” is a common pitfall that also affects scoring systems or image analysis based on two-dimensional images(26).

The cardiomyocyte nuclear volume was ∼1.9 fold greater in Fabry biopsies compared to controls. We found multiple relationships between cardiomyocyte nuclear volume and clinically relevant parameters. Thus, cardiomyocyte nuclear volume progressively increased with age and was the only statistically significant independent predictor of LVMI and directly correlated with septal thickness. Moreover, it was the only structural parameter that correlated with plasma lyso-GL3. Nuclear enlargement is a common finding during cellular hypertrophy, activation, injury or malignancy. Nuclear and cytoplasmic volumes are related to each other. A constant nucleocytoplasmic ratio has been reported in many cell types from yeasts to plants and animals(27–29). This ratio, although different among various cell types, remains more or less constant for a given cell type(30). Consistent with this notion, cardiomyocyte nuclear enlargement and cellular hypertrophy were proportional in Fabry biopsies, keeping the nucleocytoplasmic ratio similar to the control biopsies, although these results need to be confirmed in larger cohorts.

Exertional chest pain due to myocardial ischemia is common among adults with Fabry disease and is typically associated with reduced microvascular blood flow despite normal coronary angiograms(31, 32). Chimenti et al. attributed these findings to presence of remarkable luminal narrowing of intramural small arteries in myocardial biopsies(32). We found a direct correlation between the residual α-Gal-A activity and the volume fraction of blood vessels in cardiac parenchyma, suggesting that reduced microvasculature density may be an additional mechanism of myocardial ischemia and fibrosis in Fabry disease, similar to what has been reported in other conditions in the heart or other organs(33–35). Various mechanisms have been proposed to explain endothelial dysfunction in Fabry disease(36–38). Further studies to confirm our findings and explore mechanisms of microvascular rarefaction in Fabry cardiomyopathy will be necessary, especially since the microvascular blood flow impairment appears to persist despite ERT(31,32). Nevertheless, endothelial GL3 accumulation is not relevant in this case since post-ERT patients, similar to later-onset variants typically have no GL3 inclusions in their endothelial cells.

This study faced several limitations. The number of biopsies studied were limited. However, the study was confined to adult male ERT-naïve patients with Fabry disease, all with IVS4 + 919G>A GLA mutation. These together with systematic morphometric, imaging and clinical assessments led to unraveling of important findings despite this limitation. The downside would be a concern of how widely the results of this study can be extrapolated to a very heterogeneous condition, such as Fabry disease. Nevertheless, the methodology introduced in this work opens up the opportunity to perform systematic quantitative studies in future larger cohorts with a wider spectrum of Fabry phenotype. Cardiac tissues in our study were obtained from the right interventricular septum through right heart catheterization to reduce procedural risk. There may be concern that our measurements done in the right ventricular tissue are not representative for conditions in left ventricle. However, autopsy studies demonstrated that cardiac hypertrophy and GL3 accumulations are present in both ventricles in Fabry patients(39). The imaging studies by echocardiography and MRI also showed the degree of structural and functional changes in the right ventricle is mostly comparable to that in the left ventricle(40, 41). Moreover, multiple relationships between the biopsy structural parameters from the right ventricle and LVMI or septal thickness suggest that the structural parameters measured in the right ventricular biopsy are representative for evaluation of Fabry cardiomyopathy. On the other hand, although LGE in MRI was present in 9/10 patients, we did not find associations between the amount of interstitial compartment and clinical variables. A possible explanation would be that more prominent cardiomyocyte hypertrophy (relative to interstitial fibrosis) may have obscured detection of interstitial fibrosis in relative terms (reference trap). Moreover, interstitial fibrosis is a more prominent feature in the left ventricle, where LGE was detected in our patients, while the biopsies were from the right ventricle.

To summarize, this study unravels relevance of unbiased quantitative structural studies in assessment of Fabry cardiomyopathy and introduces several candidate biopsy biomarkers with potential applications as clinical trial endpoints for novel therapeutic approaches, and in better understanding the pathophysiology of the disease or for guidance of clinical decisions.

## Data Availability

The data that support the findings of this study are available on request from the corresponding authors.

## Acknowledgements

We would like to thank Mr. Edward Parker at Vision Core Research Laboratory for assisting with electron microscopy work and Dr. Aurelio Silvestroni for preparing the graphs and assisting with statistics. This work was supported by a grant (P.I. Michael Mauer and Behzad Najafian) from the National Institutes of Health Lysosomal Disease Network [(U54NS065768), a part of the NCATS Rare Diseases Clinical Research Network (RDCRN). RDCRN is an initiative of the Office of Rare Diseases Research (ORDR), NCATS, funded through a collaboration between NCATS and the National Institute of Neurological Disorders and Stroke (NINDS), and the National Institute of Diabetes and Digestive and Kidney Diseases.

## Abbreviations

α-Gal-A: α-galactosidase-A
CM: cardiomyocyte
CMN: cardiomyocyte nucleus
GL-3: globotriaosylceramide
EM: electron microscopy
ERT: enzyme replacement therapy
LV: left ventricle
LVEF: 
LVMI: Left ventricular mass index
LVH: left ventricular hypertrophy
LGE: late gadolinium enhancement
PSI: point-sampled intercept
V(CMN): volume of cardiomyocyte nuclei
Vv(CMN/CM): Volume fraction of cardiomyocyte nuclei per cardiomyocyte
Vv(Inc/CM): volume fraction of GL3 inclusions per cardiomyocyte
Vv(Ves/Heart): volume fraction of blood vessels in cardiac parenchyma
Vv(Int/Heart): volume fraction of interstitial compartment in cardiac parenchyma
V(Inc/CM): volume of GL3 inclusions per cardiomyocyte

---

**Supplemental Figure 1.**
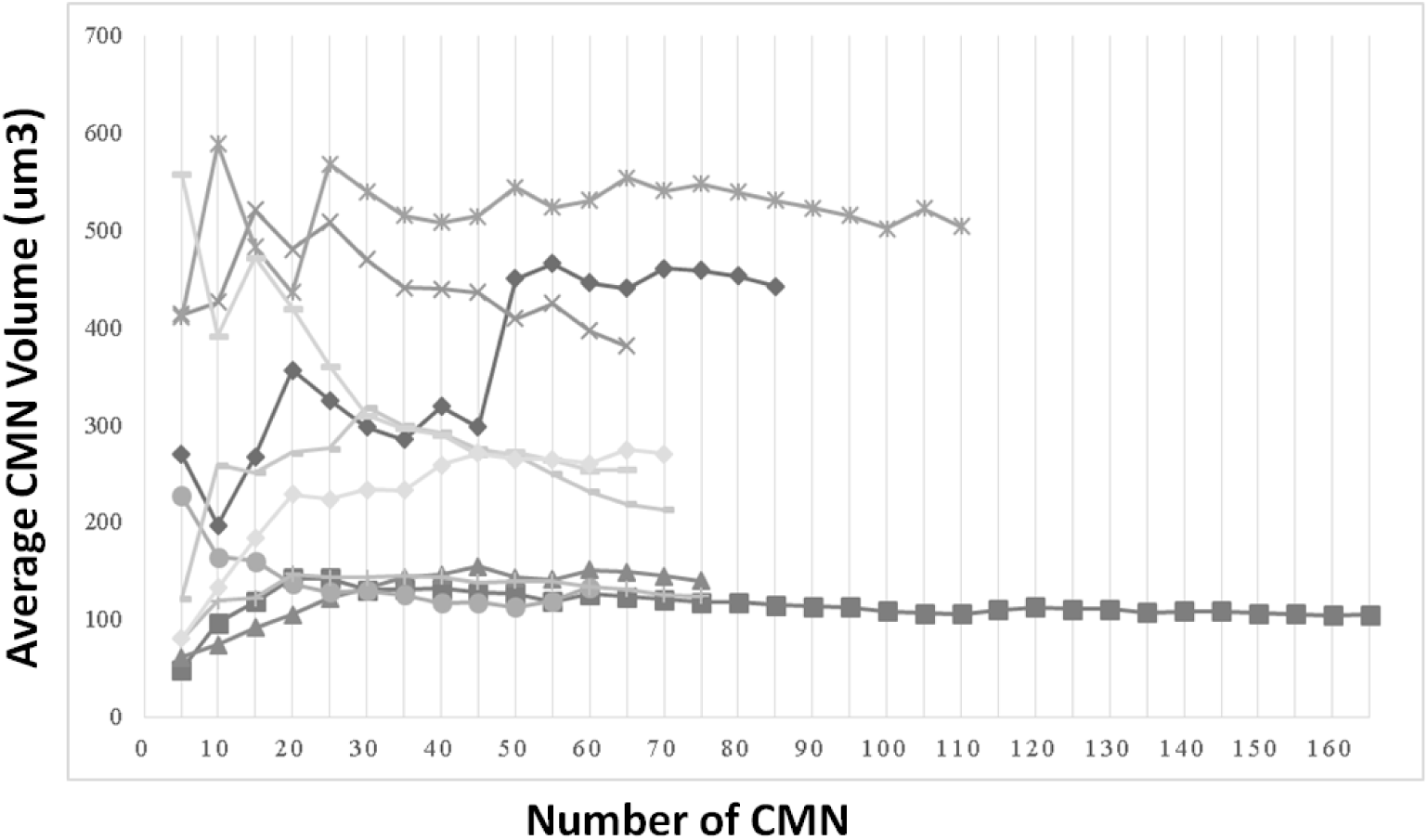
The relationship between the average cardiomyocyte nuclear volume [V(CMN)] estimated by the PSI method and incremental sampling of CMN (increment step=5). After sampling ∼60 CMN, the average V(CMN) stabilizes and inclusion of additional CMN does not noticeably change the average.

**Suppl. Figure 2.**
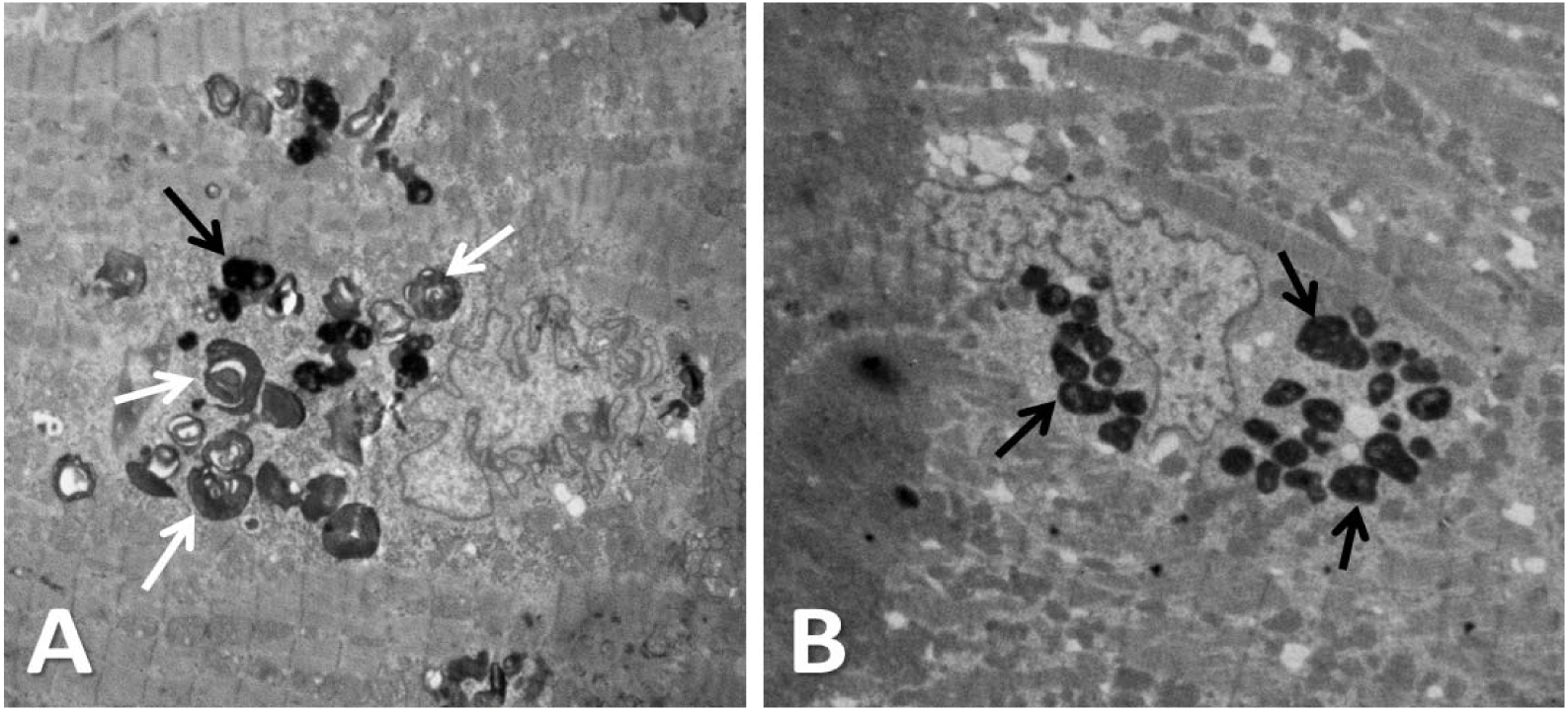
Comparison of GL3 inclusions and lipofuscin granules. **(A)** An endomyocardial biopsy from a Fabry patient shows typical multilamellar GL3 inclusion (white arrows) and a few solid electron dense round granules (dark arrows) similar to lipofuscin granules.**(B)** A control non-Fabry myocardium with lipofuscin granules (black arrows).

## References

1. Desnick RJ, Ioannou YA, Eng CM. α-galactosidase A deficiency: Fabry disease. In: Scriver CR, Beaudet AL, Sly WS, Valle D, editors. The Metabolic and Molecular Bases of Inherited Disease. New York: McGraw-Hill; 2001. p. 3733–74.

2. Germain DP. Fabry disease. Orphanet J Rare Dis. 2010;5:30.

3. Wilcox WR, Oliveira JP, Hopkin RJ, Ortiz A, Banikazemi M, Feldt-Rasmussen U, et al. Females with Fabry disease frequently have major organ involvement: lessons from the Fabry Registry. Mol Genet Metab. 2008;93(2):112–28.

4. Waldek S, Patel MR, Banikazemi M, Lemay R, Lee P. Life expectancy and cause of death in males and females with Fabry disease: findings from the Fabry Registry. Genet Med. 2009;11(11):790–6.

5. Najafian B, Tondel C, Svarstad E, Gubler MC, Oliveira JP, Mauer M. Accumulation of Globotriaosylceramide in Podocytes in Fabry Nephropathy Is Associated with Progressive Podocyte Loss. J Am Soc Nephrol. 2020;31(4):865–75.

6. Najafian B, Tondel C, Svarstad E, Sokolovkiy A, Smith K, Mauer M. One Year of Enzyme Replacement Therapy Reduces Globotriaosylceramide Inclusions in Podocytes in Male Adult Patients with Fabry Disease. PLoS One. 2016;11(4):e0152812.

7. Najafian B, Svarstad E, Bostad L, Gubler MC, Tondel C, Whitley C, et al. Progressive podocyte injury and globotriaosylceramide (GL-3) accumulation in young patients with Fabry disease. Kidney Int. 2011;79(6):663–70.

8. Thurberg BL, Fallon JT, Mitchell R, Aretz T, Gordon RE, O’Callaghan MW. Cardiac microvascular pathology in Fabry disease: evaluation of endomyocardial biopsies before and after enzyme replacement therapy. Circulation. 2009;119(19):2561–7.

9. Thurberg BL, Rennke H, Colvin RB, Dikman S, Gordon RE, Collins AB, et al. Globotriaosylceramide accumulation in the Fabry kidney is cleared from multiple cell types after enzyme replacement therapy. Kidney Int. 2002;62(6):1933–46.

10. Mauer M, Sokolovskiy A, Barth JA, Castelli JP, Williams HN, Benjamin ER, et al. Reduction of podocyte globotriaosylceramide content in adult male patients with Fabry disease with amenable GLA mutations following 6 months of migalastat treatment. J Med Genet. 2017;54(11):781–6.

11. Ramaswami U, Bichet DG, Clarke LA, Dostalova G, Fainboim A, Fellgiebel A, et al. Low-dose agalsidase beta treatment in male pediatric patients with Fabry disease: A 5-year randomized controlled trial. Mol Genet Metab. 2019;127(1):86–94.

12. Wang WT, Sung SH, Liao JN, Hsu TR, Niu DM, Yu WC. Cardiac manifestations in patients with classical or cardiac subtype of Fabry disease. J Chin Med Assoc. 2020;83(9):825–9.

13. Hsu TR, Hung SC, Chang FP, Yu WC, Sung SH, Hsu CL, et al. Later Onset Fabry Disease, Cardiac Damage Progress in Silence: Experience With a Highly Prevalent Mutation. J Am Coll Cardiol. 2016;68(23):2554–63.

14. Lin HY, Chong KW, Hsu JH, Yu HC, Shih CC, Huang CH, et al. High incidence of the cardiac variant of Fabry disease revealed by newborn screening in the Taiwan Chinese population. Circ Cardiovasc Genet. 2009;2(5):450–6.

15. Liao HC, Huang YH, Chen YJ, Kao SM, Lin HY, Huang CK, et al. Plasma globotriaosylsphingosine (lysoGb3) could be a biomarker for Fabry disease with a Chinese hotspot late-onset mutation (IVS4+919G>A). Clin Chim Acta. 2013;426:114–20.

16. Daitx VV, Mezzalira J, Goldim MP, Coelho JC. Comparison between alpha-galactosidase A activity in blood samples collected on filter paper, leukocytes and plasma. Clin Biochem. 2012;45(15):1233–8.

17. Drazner MH, Dries DL, Peshock RM, Cooper RS, Klassen C, Kazi F, et al. Left ventricular hypertrophy is more prevalent in blacks than whites in the general population: the Dallas Heart Study. Hypertension. 2005;46(1):124–9.

18. Rickers C, Wilke NM, Jerosch-Herold M, Casey SA, Panse P, Panse N, et al. Utility of cardiac magnetic resonance imaging in the diagnosis of hypertrophic cardiomyopathy. Circulation. 2005;112(6):855–61.

19. Beer M, Weidemann F, Breunig F, Knoll A, Koeppe S, Machann W, et al. Impact of enzyme replacement therapy on cardiac morphology and function and late enhancement in Fabry’s cardiomyopathy. Am J Cardiol. 2006;97(10):1515–8.

20. Kawel-Boehm N, Hetzel SJ, Ambale-Venkatesh B, Captur G, Francois CJ, Jerosch-Herold M, et al. Reference ranges ("normal values") for cardiovascular magnetic resonance (CMR) in adults and children: 2020 update. J Cardiovasc Magn Reson. 2020;22(1):87.

21. Maruyama H, Miyata K, Mikame M, Taguchi A, Guili C, Shimura M, et al. Effectiveness of plasma lyso-Gb3 as a biomarker for selecting high-risk patients with Fabry disease from multispecialty clinics for genetic analysis. Genet Med. 2019;21(1):44–52.

22. Linhart A, Kampmann C, Zamorano JL, Sunder-Plassmann G, Beck M, Mehta A, et al. Cardiac manifestations of Anderson-Fabry disease: results from the international Fabry outcome survey. Eur Heart J. 2007;28(10):1228–35.

23. Sachdev B, Takenaka T, Teraguchi H, Tei C, Lee P, McKenna WJ, et al. Prevalence of Anderson-Fabry disease in male patients with late onset hypertrophic cardiomyopathy. Circulation. 2002;105(12):1407–11.

24. Frustaci A, Chimenti C, Doheny D, Desnick RJ. Evolution of cardiac pathology in classic Fabry disease: Progressive cardiomyocyte enlargement leads to increased cell death and fibrosis, and correlates with severity of ventricular hypertrophy. Int J Cardiol. 2017.

25. Elleder M, Bradova V, Smid F, Budesinsky M, Harzer K, Kustermann-Kuhn B, et al. Cardiocyte storage and hypertrophy as a sole manifestation of Fabry’s disease. Report on a case simulating hypertrophic non-obstructive cardiomyopathy. Virchows Arch A Pathol Anat Histopathol. 1990;417(5):449–55.

26. Brown DL. Bias in image analysis and its solution: unbiased stereology. J Toxicol Pathol. 2017;30(3):183–91.

27. Jorgensen P, Edgington NP, Schneider BL, Rupes I, Tyers M, Futcher B. The size of the nucleus increases as yeast cells grow. Mol Biol Cell. 2007;18(9):3523–32.

28. Edens LJ, White KH, Jevtic P, Li X, Levy DL. Nuclear size regulation: from single cells to development and disease. Trends Cell Biol. 2013;23(4):151–9.

29. Webster M, Witkin KL, Cohen-Fix O. Sizing up the nucleus: nuclear shape, size and nuclear-envelope assembly. J Cell Sci. 2009;122(Pt 10):1477–86.

30. Gregory T. Genome size evolution in animals. In: Gregory T, editor. The evolution of the genome. London: Elsevier Academic Press; 2005. p. 4-87.

31. Elliott PM, Kindler H, Shah JS, Sachdev B, Rimoldi OE, Thaman R, et al. Coronary microvascular dysfunction in male patients with Anderson-Fabry disease and the effect of treatment with alpha galactosidase A. Heart. 2006;92(3):357–60.

32. Chimenti C, Morgante E, Tanzilli G, Mangieri E, Critelli G, Gaudio C, et al. Angina in fabry disease reflects coronary small vessel disease. Circ Heart Fail. 2008;1(3):161–9.

33. Xiao Y, Liu Y, Liu J, Kang YJ. The Association Between Myocardial Fibrosis and Depressed Capillary Density in Rat Model of Left Ventricular Hypertrophy. Cardiovasc Toxicol. 2018;18(4):304–11.

34. Sun IO, Santelli A, Abumoawad A, Eirin A, Ferguson CM, Woollard JR, et al. Loss of Renal Peritubular Capillaries in Hypertensive Patients Is Detectable by Urinary Endothelial Microparticle Levels. Hypertension. 2018;72(5):1180–8.

35. Olufsen MS, Hill NA, Vaughan GD, Sainsbury C, Johnson M. Rarefaction and blood pressure in systemic and pulmonary arteries. J Fluid Mech. 2012;705:280–305.

36. Satoh K. Globotriaosylceramide induces endothelial dysfunction in fabry disease. Arterioscler Thromb Vasc Biol. 2014;34(1):2–4.

37. Do HS, Park SW, Im I, Seo D, Yoo HW, Go H, et al. Enhanced thrombospondin-1 causes dysfunction of vascular endothelial cells derived from Fabry disease-induced pluripotent stem cells. EBioMedicine. 2020;52:102633.

38. Shu L, Vivekanandan-Giri A, Pennathur S, Smid BE, Aerts JM, Hollak CE, et al. Establishing 3-nitrotyrosine as a biomarker for the vasculopathy of Fabry disease. Kidney Int. 2014;86(1):58–66.

39. Sheppard MN, Cane P, Florio R, Kavantzas N, Close L, Shah J, et al. A detailed pathologic examination of heart tissue from three older patients with Anderson-Fabry disease on enzyme replacement therapy. Cardiovasc Pathol. 2010;19(5):293–301.

40. Palecek T, Dostalova G, Kuchynka P, Karetova D, Bultas J, Elleder M, et al. Right ventricular involvement in Fabry disease. J Am Soc Echocardiogr. 2008;21(11):1265–8.

41. Niemann M, Breunig F, Beer M, Herrmann S, Strotmann J, Hu K, et al. The right ventricle in Fabry disease: natural history and impact of enzyme replacement therapy. Heart. 2010;96(23):1915–9.

